# A Long-Lasting Sanitizing Skin Protectant based on CAGE, a Choline and Geranic Acid Eutectic

**DOI:** 10.1101/2020.08.04.20161067

**Authors:** Marina Shevachman, Abhirup Mandal, Samir Mitragotri, Nitin Joshi

## Abstract

The recent outbreak and rapid spread of coronavirus disease 2019 (COVID-19) is a global pandemic and a massive public health crisis. COVID-19 has also had a severe impact on the quality of life and mental health. While different health authorities such as WHO and CDC are encouraging adoption of strategies including hand washing and use of facemasks to reduce the spread of the pathogens and infections, adoption of these approaches requires substantial commitment. Current hand sanitizers based on ethanol provide immediate protection, however, the protection rendered by such sanitizers is very short-lived due to their rapid evaporation. A long-lasting sanitizing skin protectant that can effectively inactivate SARS-CoV-2 and provide persistent efficacy over several hours will provide people the freedom to carry on with their activities without constant concerns about the cleanliness of their hands. Herein, we describe a novel skin sanitizer, IonLAST™, based on an ionic liquid/deep eutectic solvent, formed by GRAS materials, choline and geranic acid (CAGE, CG-101), that provides protection for at least 4h after a single application. IonLAST™ was formulated as a gel that facilitates easy application on the skin. Tolerance of CG-101 was substantiated through a study in human volunteers. In vitro studies confirmed that IonLAST™ effectively inactivates a human coronavirus hCoV229E. A second human clinical study established that a single application of IonLAST™ imparts protection against microbes that lasts up to several hours.

## Introduction

Severe acute respiratory syndrome coronavirus 2 (SARS-CoV-2) has emerged as a global pandemic that has infected >13million people worldwide with 580,000 deaths reported as of July 14, 2020[1]. The zoonotic virus, SARS-CoV-2 is responsible for the coronavirus disease (COVID-19), which is associated with severe respiratory distress, fever or chills, cough, sore throat, congestion, nausea, and diarrhea[2]. Older adults and individuals with underlying health conditions such as heart or lung diseases and diabetes patients are at a higher risk of experiencing mortality from the infection[3, 4]. The World Health Organization (WHO) declared COVID-19 outbreak as a pandemic on March 11, 2020 and since then, the number of confirmed cases, deaths, and economic losses arising fromSARS-CoV-2 have amassed to an alarming level[5].

SARS-CoV-2 pandemic is not only a significant public concern in terms of human health, but its impact on the economy is also far-reaching[6]. Apart from disproportionately affecting the vulnerable, especially the elderly and minorities, it has ravaged the economic wellbeing of many, and has placed an incalculable burden on local governments. In a mere few months, it has had an impact of greater than $2T on the economy, causing the largest global recession in history[7]. In fact, COVID-19 is expected to result in long-term health issues straining the healthcare system more than ever before[8]. Even as the number of new cases seem to plateau in certain parts of the US, the recent uptick in number of new cases in other parts of the country suggests that this is not merely an issue to be managed by the healthcare system, but a call for everyone in the society to play their part to minimize the risk of transmission[9].

SARS-CoV-2 is highly contagious and can be easily transmitted from human-to-human even when the subjects are asymptomatic, presymptomatic or have mild symptoms[10]. While vaccines and antiviral drug discovery remain paramount to curb this deadly pandemic, virus containment is of utmost importance and will require behavioral changes from each individual[11, 12]. Regulatory agencies including the Center for Disease Control (CDC) and WHO have issued guidance for behavioral changes including handwashing for 20 seconds, wearing masks when in public and social distancing[13, 14]. Whilst these are simple in principle, strict compliance with these requirements is difficult to sustain. Since it is neither practical nor viable to completely lockdown all aspects of the economy or daily lives, methods that can improve compliance will contribute significantly towards the outcome.

Since 1994, the US Food and Drug Administration proposed ethanol at concentrations between 60% and 95% as safe and effective for hand sanitizers[15]. WHO recommends using formulations containing 80%v/v ethyl alcohol or 75%v/v isopropyl alcohol to inactivate pathogens effectively, including acute respiratory syndrome coronavirus (SARS-CoV), Middle East respiratory syndrome coronavirus (MERS-CoV) and most recently, SARS-CoV-2[16]. However, the biggest shortcoming of such alcohol-based formulations is that their protective effect lasts only till the alcohol evaporates i.e. 15 seconds-2 minutes. Once the alcohol evaporates there is no protection remaining on the hands leaving the person vulnerable to the very same risks of infection and transmission. Hence, development of an effective hand sanitizer that can generate long-lasting protective effects can offer significant benefits in minimizing rampant viral transmissions and maximize virus inactivation in a pandemic situation like SARS-CoV-2 outbreak[15, 17-19].

There has been no significant innovation in hand santizers from the 1960s when alcohol-based sanitizers were first introduced[20]. Here, we report a sanitizing skin protectant, IonLAST™ that contains an ionic liquid/deep eutectic solvent, choline geranate (CG-101, 5% w/w) formulated in ethanol. CG-101 is non-volatile and is hypothesized to remain on the skin long after ethanol evaporates thus protecting the skin for several hours. CG-101 has already been confirmed to demonstrate broad-spectrum antimicrobial properties against 47 ATCC strains of bacteria, virus, and fungi[21-24]. The mechanism of antimicrobial action is mediated through lipid extraction and membrane disruption[21]. We anticipated that the same properties of CG-101 may support its efficacy against SARS-CoV-2.

## Results

### Safety of CG-101 in human volunteers

Safety of CG-101 (100% concentration) was tested in a 3-week human repeat insult patch test (HRIPT) in 52 adult human volunteers. The concentration of CG-101 was 20 times higher than that used in IonLAST™. CG-101 was applied to the back of the subjects over a marked 2 x 2 cm^2^ area and covered with an occlusive hypoallergenic patch to maximize penetration. Distilled water was used as a negative control. The patches were removed by the subjects after 24h and a new one was applied by the site staff at the same exact site at the next visit. The subjects reported to the site for a total of 9 visits, 3 each week. During each visit the application area was observed by the study investigator and a new patch was applied. None of the subjects showed any skin reactions in the first 3 visits (7-days). 43 subjects did not display any irritation throughout the entire 21-day study. Between visits 4 through 9, 8 subjects experienced some skin irritation. However, there was no follow-up treatment required for any of these 8 subjects that showed irritation. There were no other adverse events reported in the study. Furthermore, upon completion of the 21-day study period, and a rest period of two weeks, CG-101 application with occlusion to a different part of the body, did not create any sensitization response at 24 and 48h.

### Superior antibacterial efficacy of IonLAST™

In order to determine the prolonged protective effects of IonLAST™ in contrast to the currently marketed products containing 70% ethyl alcohol, an in-vitro efficacy study was conducted using *E. coli* following modified ASTM E1153 methodologies. One milliliter of the test and comparator products were applied onto the pre-cleaned glass surfaces and dried for 30min. Glass slides were inoculated with 10μl of bacterial suspension at various timepoints (30min, 2h, and 4h). The test compound was chemically neutralized after an exposure duration of 5 min before transferring the samples to agar plates. The agar plates were visually compared for microbial growth after incubation for 48h at 37°C. No *E. coli* growth was observed for the IonLAST^TM^-treated groups for 30min, 2h as well as 4h which clearly indicated the persistent antimicrobial activity of IonLAST™. In contrast, almost similar *E. coli* growth was observed for the saline-treated group and the comparator, ethyl alcohol 70% hand sanitizer treated group (**Fig. 1**). This indicates that alcohol-based products might have limited ability to provide long lasting protective effects against microbes. On the contrary, IonLAST™ provided clear indication of long-lasting protection against pathogens and thus potentially reducing disease transmission during a pandemic.

**Fig. 1.**
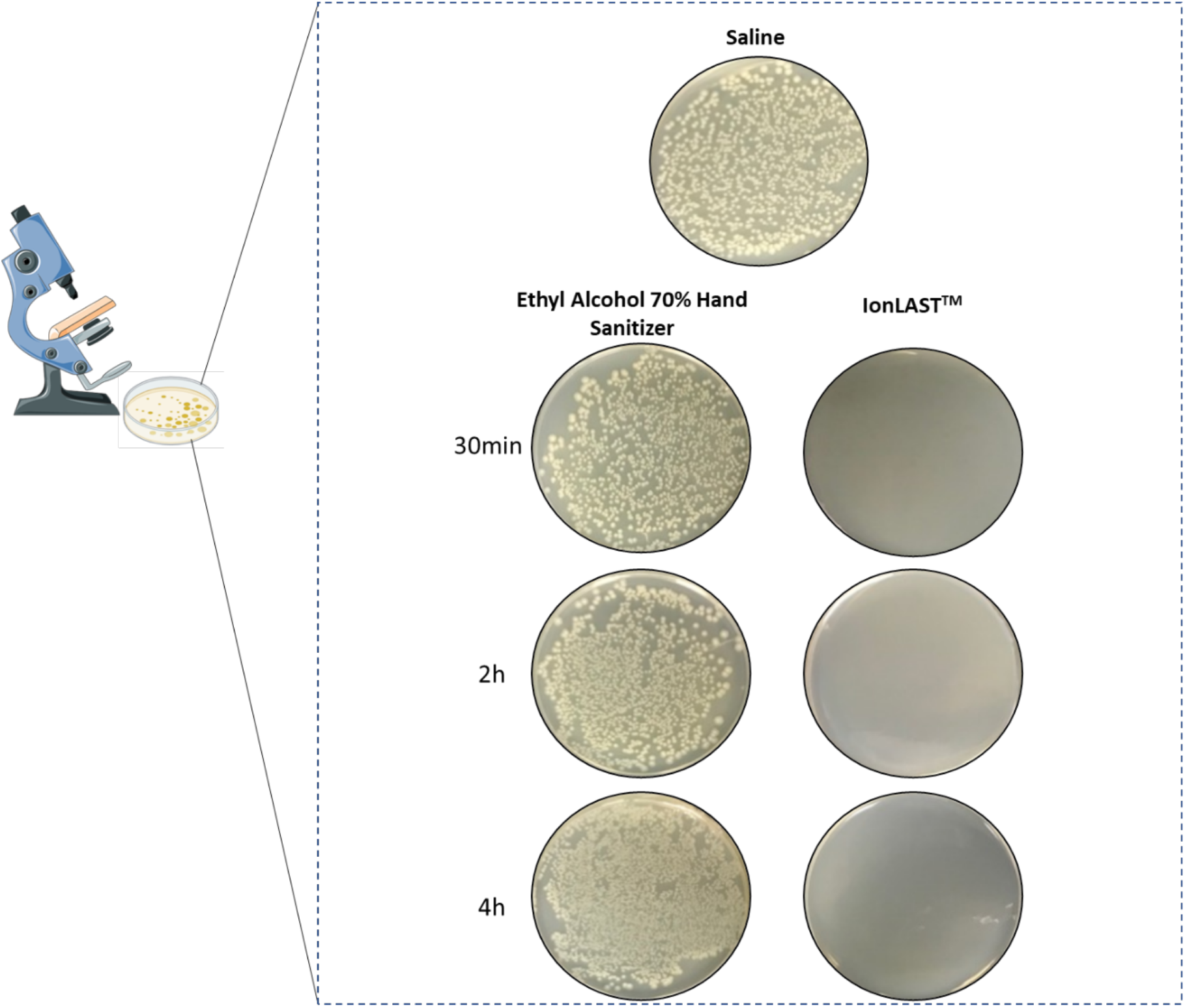
In-vitro antibacterial efficacy against *E. coli*. Number of viable test microorganisms treated with the test substances, ethyl alcohol 70% hand sanitizer (comparator) and IonLAST™ in comparison to the saline-treated group (negative control) The treated groups were incubated with the test substances for 30min, 2h and 4h and subsequently exposed to *E Coli* for 5min and then neutralized. Bacterial growth was assessed on agar plates after a 48-hour incubation period.

### Protection against hCoV229E

Based on the previously demonstrated broad-spectrum antimicrobial properties of CG-101[23], we anticipated that CG-101 and the skin sanitizer containing CG-101 (IonLAST™) would effectively deactivate coronavirus likely through disruption of the phospholipid bilayer glycoproteinaceous envelope. We evaluated the virucidal effects of IonLAST™ gel and the active CG-101 (5% w/w in purified water) against human Coronavirus strain 229E (hCoV229E) using a virucidal suspension test (in-vitro time-kill method) based on industry/regulatory-relevant global standardized methodologies (ASTM E1052-20). The log_10_ reductions from the initial population of the hCoV229E following 15s and 30s exposure to IonLAST™ was found to be >4.00. In fact, the percent reduction for both the time points was >99.99. Similarly, we evaluated the virucidal activity of CG-101 (5% w/w) using the quantitative suspension test for exposure times, 5min and 10min. The percent and log_10_ reductions from the initial population of the hCoV229E following 5min and 10min exposure to CG-101 (5% w/w) was found to be >99.99 and >4.00 respectively (**Fig. 2 and Tables S1 and 2**).

**Fig. 2.**
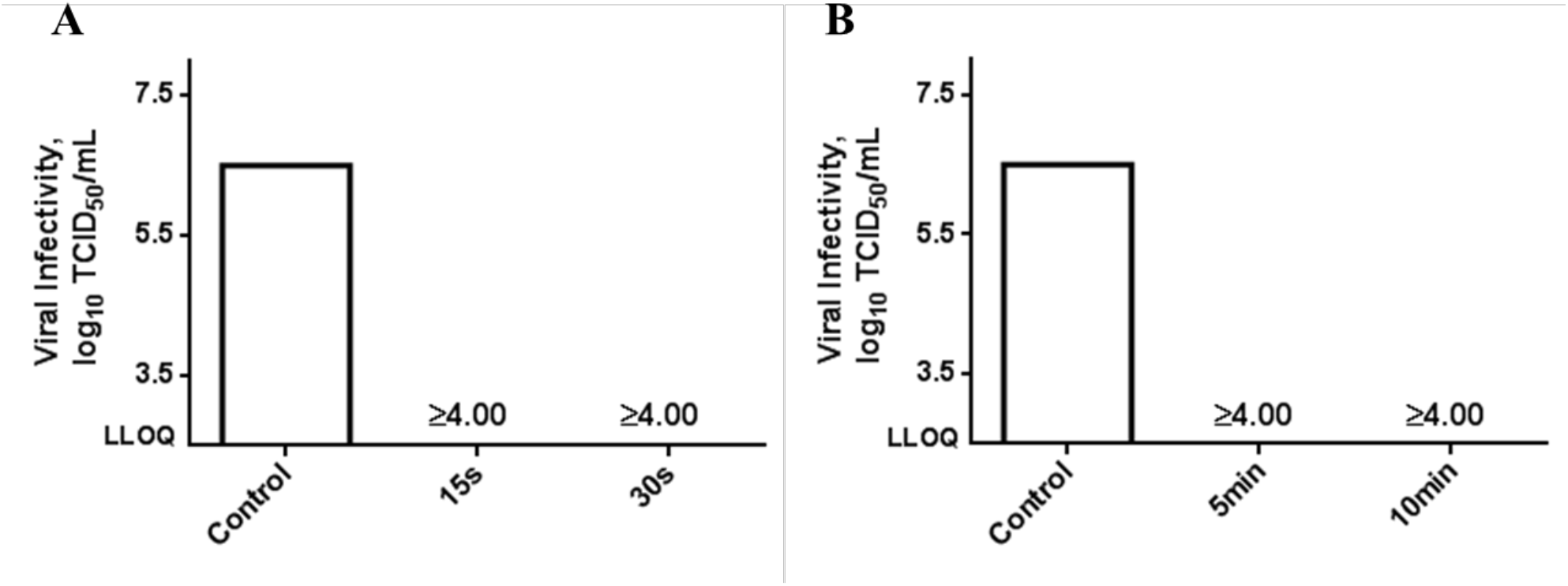
GLP virucidal efficacy test against human coronavirus strain 229E (hCoV229E) Virucidal infectivity to MRC-5 cells for (A) IonLAST™ and (B) CG-101 (5% w/w) treated viral suspensions in comparison to virus control (untreated) for different exposure times. Log_10_ reduction are included above the bar. Viral titers are displayed as TCID_50_/mL values (n=4). LLOQ, lower limit of quantification; TCID_50_/mL, 50% tissue culture infectious dose.

### Prolonged residual antimicrobial efficacy of IonLAST™ in humans

Clinical studies approved by an IRB were performed at Bioscience laboratories, Inc. (testing facility) in compliance with good clinical practice regulations to test the effectiveness of IonLAST™ to provide extended protection against microbes. Specifically, IonLAST™ was applied to the forearms of human volunteers and the randomly assigned test sites on each forearm were challenged by application of *Staphylococcus aureus (S. aureus)* at various times after the application of IonLAST™. The residual antimicrobial efficacy of IonLAST™ was tested using a modification of the standardized test method described in ASTM E2752-10 (2015) in 12 healthy subjects. A significant (≥5.15) log_10_ reduction in *S. aureus* from the control was observed following 30min., 2h and 4h post-IonLAST™ application.

Immediately following application and 30-min air-dry of the IonLAST™, the mean log_10_ microbial recovery for the treated subjects was found to be 0.86±0.00 in contrast to 6.30±0.05 for the untreated subjects. Furthermore, the log_10_ microbial recovery, 2h following application of the IonLAST™, was found to be 0.86±0.00 for the treated subjects vs 6.24±0.07 for untreated subjects. Interestingly, post 4h application of IonLAST™, the log_10_ microbial recovery for treated subjects was also 0.86±0.00 in comparison to 6.24±0.12 for untreated subjects. Note that the lowest detectable limit of the study was 0.86 log_10_ CFU/cm^2^. IonLAST™ thus demonstrated prolonged microbial protection for up to 4h after a single application (**Fig. 3 and Table S3**). These data and the lack of any trend from 0.5h, 2h and 4h suggest that the effectiveness of IonLAST™ could persist beyond 4h.

**Fig. 3.**
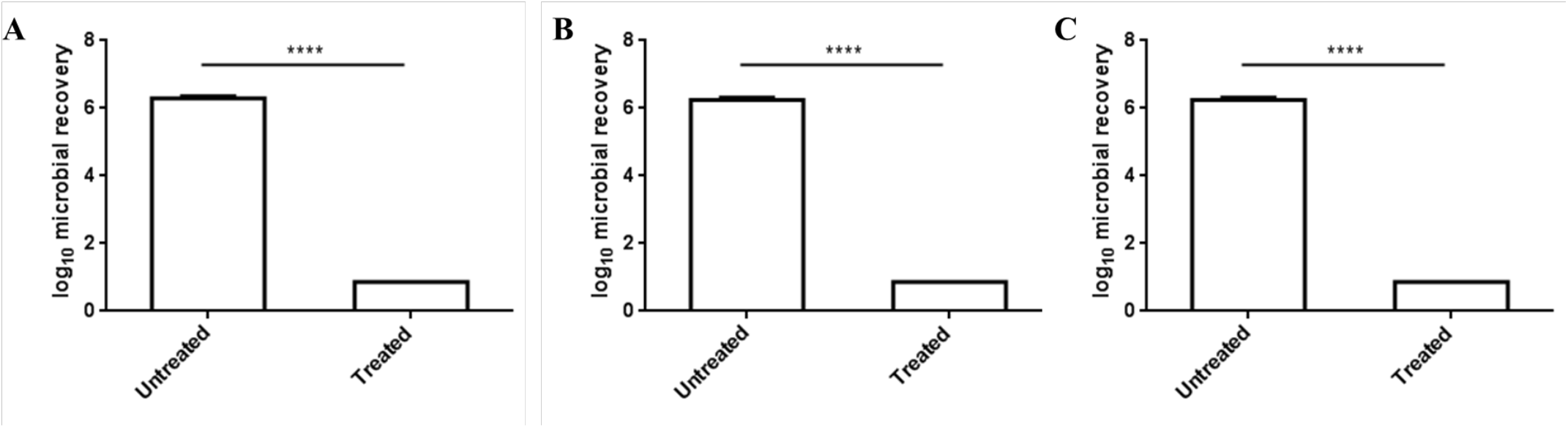
Residual antimicrobial efficacy against *S. aureus*. The means of log_10_ microbial recovery in comparison to untreated subjects (A) immediately (30min), (B) 2h, and (C) 4h following application of the Test Product: IonLAST™. Data are averages ± SEM, statistics by two-way ANOVA with Bonferroni’s multiple comparison-test. ****P < 0.0001 (n=11).

## Discussion

SAR-CoV-2, the virus that causes the COVID-19 disease has evolved into a global pandemic and is highly contagious affecting >25million people worldwide over a span of just 6 months. While the development of effective vaccines and antiviral therapies remain at the forefront of the global efforts to curb this deadly infection, the immediate response plan to control COVID-19 should include breaking the chain of transmission of the virus and preventing associated illness and death[25]. Current evidence suggests that SARS-CoV-2 predominantly spreads through person-to-person via saliva and respiratory secretions or droplets[26]. Other modes of transmission could be contaminated objects or surfaces (fomite transmission), fecal-oral, bloodborne, mother-to-child, and animal-to-human transmission[27]. The relative roles of various routes are unclear and will require additional epidemiological data as well as mechanistic information about the virus entry. For example, angiotensin-converting enzyme 2 (ACE2), the cell receptor for SARS-CoV-2 entry is abundantly present in blood vessels/capillaries of the skin, the basal layer of the epidermis, and hair follicles[28, 29].

Hand hygiene is widely recognized for playing a major role in limiting the spread and transmission of such infectious diseases[18]. In fact, in order to cope with such a pandemic situation, CDC has specifically recommended washing of hands with soap and water whenever possible. However, it has been reported that frequent washing hands with water and soap may damage skin potentially leading to dermatitis[30]. While alcohol-based hand sanitizers with at least 60% alcohol can help in avoiding sickness and spreading germs to others, unfortunately the protective effect of alcohol lasts only till the alcohol evaporates i.e. less than a couple of minutes. Frequent handwashing is also burdensome and disruptive in many situations, and in some cases just not practical[31].

Inspired by the fact, that CG-101 exhibits broad-spectrum antimicrobial properties against a range of bacteria, virus, and fungi, we developed an alcohol-based sanitizing skin protectant containing CG-101 as a countermeasure to restrict the rampant spread of the COVID-19 infection. CG-101 inactivates pathogens by extracting lipids and disrupting the cell membrane[21]. More importantly, unlike ethanol, which is highly volatile, CG-101, being an ionic liquid/deep eutectic solvent, has a very low vapor pressure and can remain on the skin long after ethanol evaporates. This long-lasting residence of CG-101 is expected to be responsible for long-lasting protection against SARS-CoV-2 thus limiting the need for frequent hand washing.

Human volunteer studies confirmed the safety of the active, CG-101(~100% concentration). HRIPT, an industry standard test for evaluating the potential of a test material to induce sensitization in humans after repeated exposure was adopted[32]. Among 52 adult human volunteers who were treated with 20x higher CG-101 than that used in the IonLAST™, 43 subjects did not display any irritation throughout the entire 21-day study. There were no other adverse events reported in the study. Furthermore, following completion of the study and a rest period of two weeks, CG-101 application to a different part of the body, did not create any sensitization response at 24 and 48 h which indicates the potential of CG-101 as a safe additive for topical products.

In-vitro bactericidal assay was conducted to compare the longevity of antibacterial efficacy of IonLAST™ in contrast to currently marketed alcohol-based products against *E. coli*. The visual difference in the number of viable test microorganisms for the alcohol-based product and IonLAST™ was striking for all the time points including 30min, 2h and 4h. No notable difference in the microbial growth was observed between the untreated and alcohol-based hand sanitizer treated groups (**Fig. 1**) indicating the short duration of protection rendered by ethanol. This signifies the persistence of protective effects of IonLAST™ against *E. coli* and possibly other pathogens for up to 4h.

The encouraging results obtained from the bactericidal efficacy studies, motivated us to test IonLAST™ against hCoV229E. As theorized, CG-101 was able to deactivate the virus, generating excellent virucidal efficacy against hCoV229E. IonLAST™ generated >4.00 log_10_ reductions in viral titers following 15s and 30s exposure. Moreover, virucidal activity of the active, CG-101 (5% w/w) for exposure times, 5min and 10min generated similar log_10_ reductions (>4.00) from the initial population of the hCoV229E as IonLAST™ (Fig. 2) indicating hCoV229E is highly susceptible to IonLAST^TM^.

Residual antimicrobial efficacy study was undertaken to measure the relative persistence of antibacterial activity under controlled clinical test conditions. The results demonstrated a significant (≥5.15) log_10_ reduction in *S. aureus* following immediate, 2h and 4h post-IonLAST™ application (**Fig. 3**). These results further support persistent pathogen inactivation efficacy of IonLAST™. Upon further clinical studies demonstrating the prolonged protection rendered against hCoV229E, IonLAST™ may open new opportunities in the recommended hand hygiene protocols for preventing disease transmission. With its long-lasting effect, IonLAST offers a promising complement to the existing CDC/WHO guidelines for good hand hygiene.

## Materials and Methods

### Clinical safety study of CG-101

For the HRIPT, skin irritation and sensitization potential of CG-101 was evaluated in a 21-day human repeat insult patch test in 52 volunteers. This cosmetic study was approved by an IRB and conducted by AMA Laboratories Inc. in New York. CG-101 (100%, liquid) was applied on the back of volunteers over a 2 x 2 cm^2^ area on each of 9 visits over a period of 3 weeks. The application area was occluded with hypoallergenic tape. The subjects were required to keep the tape for 24h. At each visit, the application site was evaluated for skin irritation on a 5-point scale (0-4). Any subjects that showed irritation rated at 3 or higher were withdrawn from the study and monitored for changes. After completion of the 3-week test period the subjects were given a 10 to 14-day rest, after which CG-101 solution was applied to a previously unexposed site. The subjects were then evaluated for any skin reactions after 24 and 48h.

### In-vitro bactericidal efficacy

In order to determine the relative bactericidal efficacy of IonLAST™ in contrast to an alcohol-based product, ASTM E1153, standard test method for efficacy of sanitizers recommended for inanimate non-food contact surfaces was modified and employed accordingly. Briefly, pre-cleaned surfaces were transferred into sterile petri plates using sterile forceps and covered with 1ml of the test substances and held for 30min at ambient temperature to dry. The inoculation with 10 μl of *E. coli* suspension was performed at 30 min, 2h and at 4h. After each inoculation, the exposure time was 5 min. After 5 min, the test compound was neutralized, and the viable bacteria were resuspended. A neutralization study was conducted to assure that the neutralizers used for the study quench the antimicrobial activity of each test material and were not toxic to the challenge species. After neutralization, the samples were plated on agar plates and transferred into the incubator set at 37°C for 48h. An average of at least ≥10^4^ CFU of bacteria were recovered from the negative and neutralization controls. The number of viable organisms on the agar plates treated with IonLAST™ were then compared visually to a negative control (untreated) and the comparator (ethyl alcohol 70% hand sanitizer) following 48 h incubation.

### Virucidal suspension test

The virucidal test was based upon ASTM E1052-20, standard practice to assess the activity of microbicides against viruses in suspension. All testing was performed in accordance with Good Laboratory Practices (GLP), as specified in 21 CFR Part 58 at BioScience Laboratories, Inc. (BSL), Bozeman, Montana. The characterization of the identity, strength, purity, composition, stability, and solubility of the test product(s) was performed by CAGE Bio Inc. The percent and log_10_ reductions from the initial population of the viral strain(s) was determined following exposure to the test product, IonLAST™, for 15s and 30s, and to the test product, CG-101 (5% w/w), for 5minand 10min. Testing was performed in 1 replicate. Plating was performed in four replicates. Coronavirus (alphacoronavirus) strain 229E (ATCC #VR-740) and MRC-5 (ATCC #CCL-171; human lung fibroblast cells) were used for the study.

### Residual antimicrobial efficacy test

The residual antimicrobial efficacy testing was carried out using a modification of the standardized test method described in ASTM E2752-10 (2015), standard guide for evaluation of residual effectiveness of antibacterial personal cleansing products. The study was conducted in compliance with Good Clinical Practice Regulations, Good Laboratory Practice Regulations, the standard operating procedures of BioScience Laboratories, Inc., the study protocol, and any protocol amendments. Bacterial recoveries were assayed after application of test material, using the forearms as a substrate. At least twelve test subjects (aged 18-65 years), with healthy skin were used in this study, and the test product was applied on one arm, and had the other arm untreated as the negative control. The test sites on both forearms were inoculated with suspensions containing *S. aureus* (ATCC #6538), immediately after the 30-minute product drying time, and at approximately 2h and 4h following test material applications. The test sites were sampled using the cup scrub procedure approximately 20min following each inoculation. The log 10 microbial recoveries of treated versus untreated sites were the basis for assessing the residual antimicrobial effectiveness of the test product.

A neutralization study was also performed to assure that the neutralizers used in the recovery medium quench the antimicrobial activity of each test material and were not toxic to the challenge species. Study procedures were based on ASTM E 1054-08(2013), standard test methods for evaluation of inactivators of antimicrobial agents. *S. aureus* (ATCC #6538) was used as the challenge species in the neutralization study.

### Statistical Analysis

For the virucidal suspension test, the Quantal test (Spearman-Kärber Method) was applied to calculate virus titer. No control of bias was performed. The MiniTab® Version 18 statistical computer package was used for all statistical calculations for the antimicrobial efficacy test. A blocked, one-factor Analysis of Variance (ANOVA) model was used:

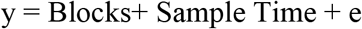

where:

y = Log_10_ Reduction = Untreated Log_10_ Recovery - Treated Log_10_ Recovery Blocks = One subject will use both configurations.

Sample Time

1, if immediate

2, if 2h 3, if 4h

e =Error Term

## Data Availability

Any data associated with this study are available from the authors upon reasonable request.

## Supplementary Materials

### Supplementary Materials and Methods

Table S1. Log_10_ reduction and percent reduction from control of hCoV229E (ATCC #VR-740) following 15- and 30s by the test product: IonLAST™

Table S2. Log_10_ reduction and percent reduction from control of hCoV229E (ATCC #VR-740) following 5- and 10min by the test product: CG-101 (5% w/w)

Table S3. Log_10_ microbial recoveries and Log_10_ microbial reductions from control of *S. aureus* (ATCC #6538), by subject; immediately (30min), 2h, and 4h following application of the test product: IonLAST™

## Author contributions

MS led characterization, formulation development; MS, AM and NJ designed the GLP antibacterial and vircucidal studies, and data analysis; MS and NJ participated in characterization, design of clinical safety study and data analysis; AM performed data summary, AM and SM summarized findings and wrote the manuscript with the help of all authors.

## Data and materials availability

Any data associated with this study are available from the authors upon reasonable request.

